# COVID-19 Epidemic Forecast in Different States of India using SIR Model

**DOI:** 10.1101/2020.05.14.20101725

**Authors:** Mukesh Jakhar, P. K. Ahluwalia, Ashok Kumar

## Abstract

(May 14, 2020)

The epidemiological data up to 12^th^ May 2020 for India and its 24 states has been used to predict COVID-19 outbreak within classical SIR (Susceptible-Infected-Recovered) model. The basic reproduction number R_0_ of India is calculated to be 1.15, whereas for various states it ranges from 1.03 in Uttarakhand to 7.92 in Bihar. The statistical parameters for most of the states indicates the high significance of the predicted results. It is estimated that the epidemic curve flattening in India will start from the first week of July and epidemic may end in the third week of October with final epidemic size ∼1,75,000. The epidemic in Kerala is in final phase and is expected to end by first week of June. Among Indian states, Maharashtra is severely affected where the ending phase of epidemic may occur in the second week of September with epidemic size of ∼55,000. The model indicates that the fast growth of infection in Punjab is from 27^th^ April 2020 to 2^nd^ June 2020, thereafter, curve flattening will start and the epidemic is expected to finished by the first week of July with the estimated number of ∼3300 infected people. The epidemic size of COVID-19 outbreak in Delhi, West Bengal, Gujrat, Tamil Nadu and Odisha can reach as large as 24,000, 18,000, 16,000, 13,000 and 11,000, respectively, however, these estimations may be invalid if large fluctuation of data occurs in coming days.

## Introduction

COVID-19 (Coronavirus disease 2019) is a disease caused by a novel virus called SARSCoV-2 (severe acute respiratory syndrome coronavirus 2) which has spread over more than 200 countries infecting more than 40 lakh peoples worldwide (as on May 12,2020) [1]. The outbreak of this disease was announced by World Health Organization (WHO) after one month from the reporting of first case on Dec. 31, 2019, in Wuhan, China and later as a pandemic on March 11, 2020 [2]. India is also in the thick of this highly infectious viral disease with over 70,000 people caught in its tentacles (as on May 12, 2020) [3].

The symptoms of COVID-19 range from mild to severe, which are indicated by mainly fever, cough, and respiratory distress. The two most important modes of transmission of corona virus are respiratory droplets and contact transmission (contaminated hands) with an incubation period 2–14 days [4–5]. The virus spread rapidly around the world and several large-size clusters of the spread have been observed worldwide including outbreaks in China, USA, Spain, Russia, UK, Italy and India. Despite strong interventions including country wide lockdowns by governments of many countries, this pandemic is nowhere near full control in most of the countries except China and South Korea.

In the absence of a COVID 19 vaccine at the moment the only accepted way to attenuate the growth is to practice good hand hygiene, using masks compulsorily and social distancing. In an effort to contain this epidemic’s spread in India, the Indian Prime Minister also announced a nation-wide lockdown from the midnight of 24th March 2020 and subsequent series of lockdowns to prevent spreading of the virus from human-to-human transmission. Since these measures have brought huge pressure on economy, it is not only important to contain the spread of the Coronavirus but also to have quantitative estimates of the spread or its abetment to estimate its impact and to plan economic and health policies to reduce the shock on economy.

Indian Government has been proactive since the end of January 2020, when the first case of infected person was detected in Kerala. Indian government had taken preventive measures well in time by announcing nationwide lockdown on 22^nd^ March 2020 and its 4^th^ phase, Lockdown 4.0, is going to start from 18^th^ of May 2020. Detailed measures [3] such as screening of passengers at airport, restricting public gatherings, suspension of transport including flights, trains and buses, increasing quarantine facility, dedicated COVID-19 hospitals, increasing sample testing etc. were taken by the government during Lockdown period. To revive the economy, Prime minister Modi has announced a package of Rs. 20 lakh crores (20 trillion) on 12^th^ May 2020.

Many studies have been recently reported by researchers to understand the dynamics of this pandemic [6–11]. The forecast of COVID-19 in the context of India has been investigated by many researchers using mathematical [12–16] and epidemiological [17–19] models but have limited studies [20–21] of individual states. Looking at the demographical and geographical diversity in India, a separate state-wise study of COVID-19 epidemic is the need of the hour.

In this paper, we present a study of epidemiological Susceptible-Infected-Recovered (SIR) model [22] for the spread of COVID-19 in various states of India. It is worth mentioning here that the forecasts using this model are as good as the quality of data available and, therefore, the progress of the spread of the virus may also affect the predictions [23].

## Methods

The SIR model is one of the simplest compartmental models which consists of three-compartment levels: Susceptible (S), Infectious (I) and Removed (R)[22]. S represents those peoples who have no immunity to the disease but they are not infectious and can be represented by the entire population, I are those who have already contracted the disease, and the R represents the recovered and diseased peoples. Also, it assumes that within the outbreak period, no significance population change takes place (e.g., through new births, deaths, migration etc.) and N = S + I + R = Constant. The SIR model can be expressed by the following set of ordinary differential equations [24]:

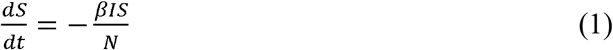

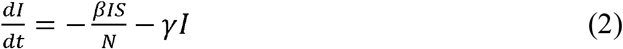

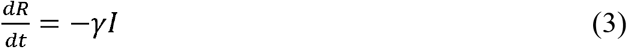

Where, is time, is the number of susceptible persons at time *t, I* = *I*(*t*), is the number of infected persons at time *t, R*(*t*), is the number of recovered persons in time *t, β* is the contact rate, and 1/*γ* is the average infectious period. In particular, the model with three equations can be reduced to one function about the total infection count (*C* = *I* + *R*) [25]. In our study, the epidemiological data has been collected from official website of Ministry of Health and Family Welfare, Government of India (https://www.mohfw.gov.in) till 12^th^ May 2020 from the date of first case detected in each state. The simulations of SIR model are performed using *fminsearch* and *ode45 functions of* MATLAB as implemented by M. Batista in references [26–27].

## Results and Discussion

The cumulative number of infected peoples in the states under consideration varies from ∼50 to ∼25,000. Maharashtra has recorded highest number of infected peoples (24427) till 12^th^May 2020, followed by 8904 in Gujrat, 8718 in Tamil Nadu and 7639 in Delhi. The states like Rajasthan, Madhya Pradesh, Uttar Pradesh, West Bengal, Punjab and Andhra Pradesh have number of cases in the range of 1000–5000 on 12^th^ May 2020 whereas rest of the states have less than 1000 cases. Kerala is the only state in India, which seems to be in total control over COVID-19 epidemic.

The results of the calculation are shown in Table 1 and in Figures 1–5. We consider different states in descending order of the number of cumulative infected cases. For each state we have calculated R_0_ (basic reproduction number), β (Average contact frequency), γ (average removal frequency), C_{end}_ (epidemic size),and S_{end}_(final number of susceptible individuals left) with the four periods of infection i.e. (i) start of acceleration, (ii) start of steady growth, (iii) start of ending phase and (iv) end of epidemic (1 case). In the figure for each state, we present two graphs: one is total number of novel Corona cases per day and second is the different epidemic phases i.e. initial exponential growth, fast growth, asymptotic slow growth and curve flattening. The statistical parameters such as coefficient of determination (R^2^), adjusted R^2^, p-value, root mean square error (RMSE) and F-statistics vs zero model for each state are listed in Table 2. The coefficient of determination (R^2^) and p-value of the model for most of the states are close to 1 and 0, respectively, indicating high statistical significance of the results.

**Table 1:** The various parameters calculated using SIR model. R_0_, β, γ, C_{end}_ and S_{end}_ represent basic reproduction number, average contact frequency, average removal frequency, epidemic size and final number of susceptible individuals left.

| India and State | $R_0$ | $\beta$ | $\gamma$ | $C_{\text{end}}$ | $S_{\text{end}}$ | Outbreak (year-2020) | Start of acceleration (year-2020) | Start of steady growth (year-2020) | Start of ending phase (year-2020) | End of epidemic (1 case) (year-2020) |
| --- | --- | --- | --- | --- | --- | --- | --- | --- | --- | --- |
| India | 1.15 | 0.66 | 0.573 | 176126 | 523659 | 03-Mar | 23-Apr | 10-Jun | 04-Jul | 16-Oct |
| Maharashtra | 1.11 | 1.008 | 0.908 | 55631 | 234823 | 11-Mar | 25-Apr | 05-Jun | 26-Jun | 09-Sep |
| Gujrat | 1.58 | 0.282 | 0.179 | 16409 | 9558 | 20-Mar | 19-Apr | 01-Jun | 22-Jun | 13-Sep |
| Tamil Nadu | 3.1 | 0.371 | 0.12 | 12555 | 701 | 18-Mar | 03-May | 20-May | 29-May | 29-Jul |
| Delhi | 1.19 | 0.486 | 0.409 | 23635 | 55146 | 03-Mar | 26-Apr | 19-Jun | 16-Jul | 15-Oct |
| Rajasthan | 1.72 | 0.269 | 0.156 | 4900 | 2080 | 03-Mar | 09-Apr | 19-May | 08-Jun | 13-Aug |
| Madhya Pradesh | 1.68 | 0.347 | 0.206 | 4053 | 1866 | 22-Mar | 09-Apr | 11-May | 27-May | 16-Jul |
| Uttar Pradesh | 1.77 | 0.273 | 0.154 | 4531 | 1748 | 04-Mar | 11-Apr | 19-May | 07-Jun | 10-Aug |
| West Bengal | 1.96 | 0.18 | 0.092 | 17522 | 4751 | 19-Mar | 11-May | 01-Jul | 26-Jul | 24-Nov |
| Andhra Pradesh | 1.12 | 0.979 | 0.874 | 2500 | 9462 | 19-Mar | 09-Apr | 17-May | 05-Jun | 15-Jul |
| Punjab | 1.28 | 0.797 | 0.621 | 3237 | 4708 | 18-Mar | 27-Apr | 21-May | 02-Jun | 01-Jul |
| Telangana | 2.44 | 0.298 | 0.122 | 1180 | 153 | 14-Mar | 31-Mar | 26-Apr | 09-May | 22-Jun |
| Jammu and Kashmir | 1.41 | 0.275 | 0.195 | 1330 | 1211 | 13-Mar | 06-Apr | 28-May | 24-Jun | 19-Aug |
| Karnataka | 1.09 | 0.789 | 0.723 | 1216 | 6031 | 12-Mar | 03-Apr | 30-May | 28-Jun | 16-Aug |
| Bihar | 7.92 | 0.18 | 0.023 | 823 | 0 | 23-Mar | 18-Apr | 15-May | 28-May | 17-Jul |
| Kerala | 2.55 | 0.271 | 0.106 | 494 | 55 | 05-Mar | 19-Mar | 16-Apr | 30-Apr | 06-Jun |
| Odisha | 1.74 | 0.19 | 0.109 | 10921 | 4449 | 21-Mar | 28-May | 22-Jul | 19-Aug | 05-Dec |
| Jharkhand | 1.86 | 0.298 | 0.16 | 191 | 61 | 02-Apr | 14-Apr | 16-May | 31-May | 22-Jun |
| Tripura | 1.87 | 2.101 | 1.123 | 148 | 47 | 10-Apr | 06-May | 11-May | 13-May | 16-May |
| Uttarakhand | 1.03 | 2.264 | 2.204 | 71 | 1071 | 20-Mar | 24-Mar | 06-May | 27-May | 04-Jun |
| Assam | 3.44 | 0.969 | 0.281 | 41 | 1 | 01-Apr | 03-Apr | 09-Apr | 12-Apr | 14-Apr |
| Himachal Pradesh | 3.23 | 0.696 | 0.215 | 36 | 2 | 22-Mar | 04-Apr | 13-Apr | 18-Apr | 20-Apr |
| Chhattisgarh | 1.23 | 0.259 | 0.211 | 83 | 132 | 24-Mar | 26-Mar | 25-May | 24-Jun | 14-Jul |

**Figure 1:**
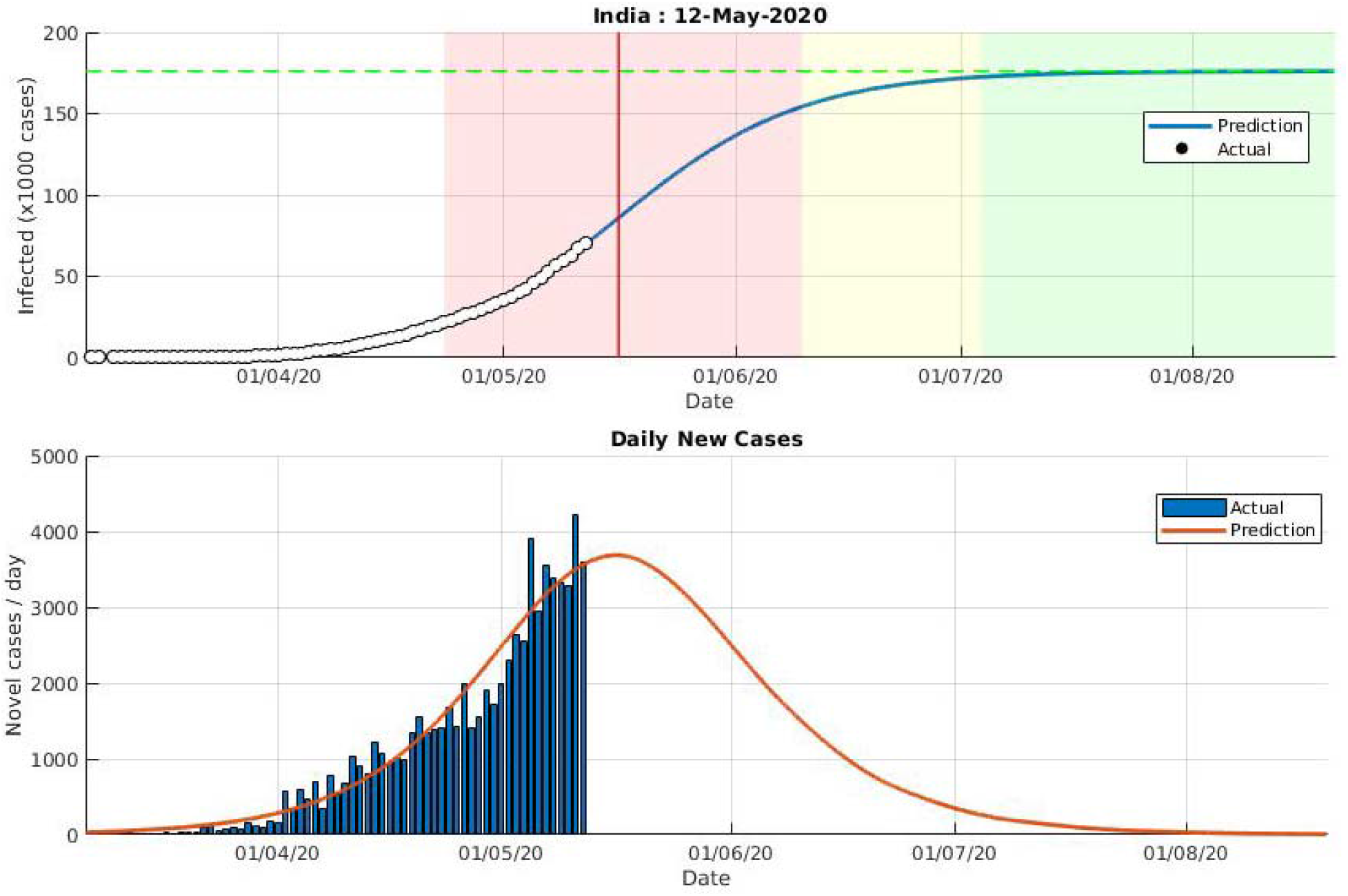
Predictions using SIR model for India using data upto 12^th^ May 2020. The different epidemic phases are shown with white, red, yellow and green colors which represent initial exponential growth, fast growth (with positive and negative phase separated by red vertical line), asymptotic slow growth and curve flattening, respectively.

**Figure 2:**
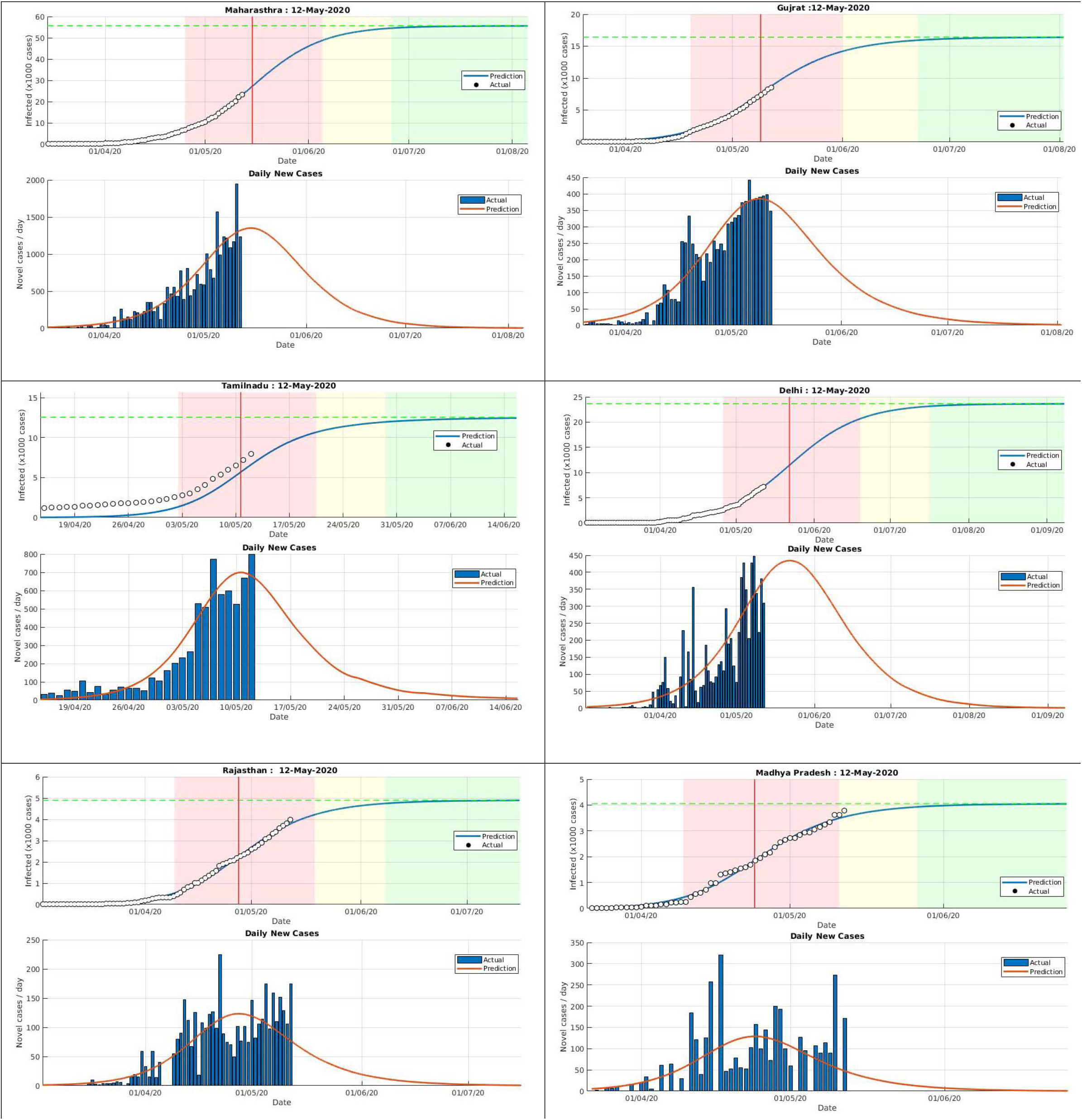
Predictions for various states (Maharashtra, Gujrat, Tamil Nadu, Delhi, Rajsthan and Madhya Pradesh) using SIR model. The different epidemic phases are shown with white, red, yellow and green colors which represent initial exponential growth, fast growth (with positive and negative phase separated by red vertical line), asymptotic slow growth and curve flattening, respectively.

**Figure 3:**
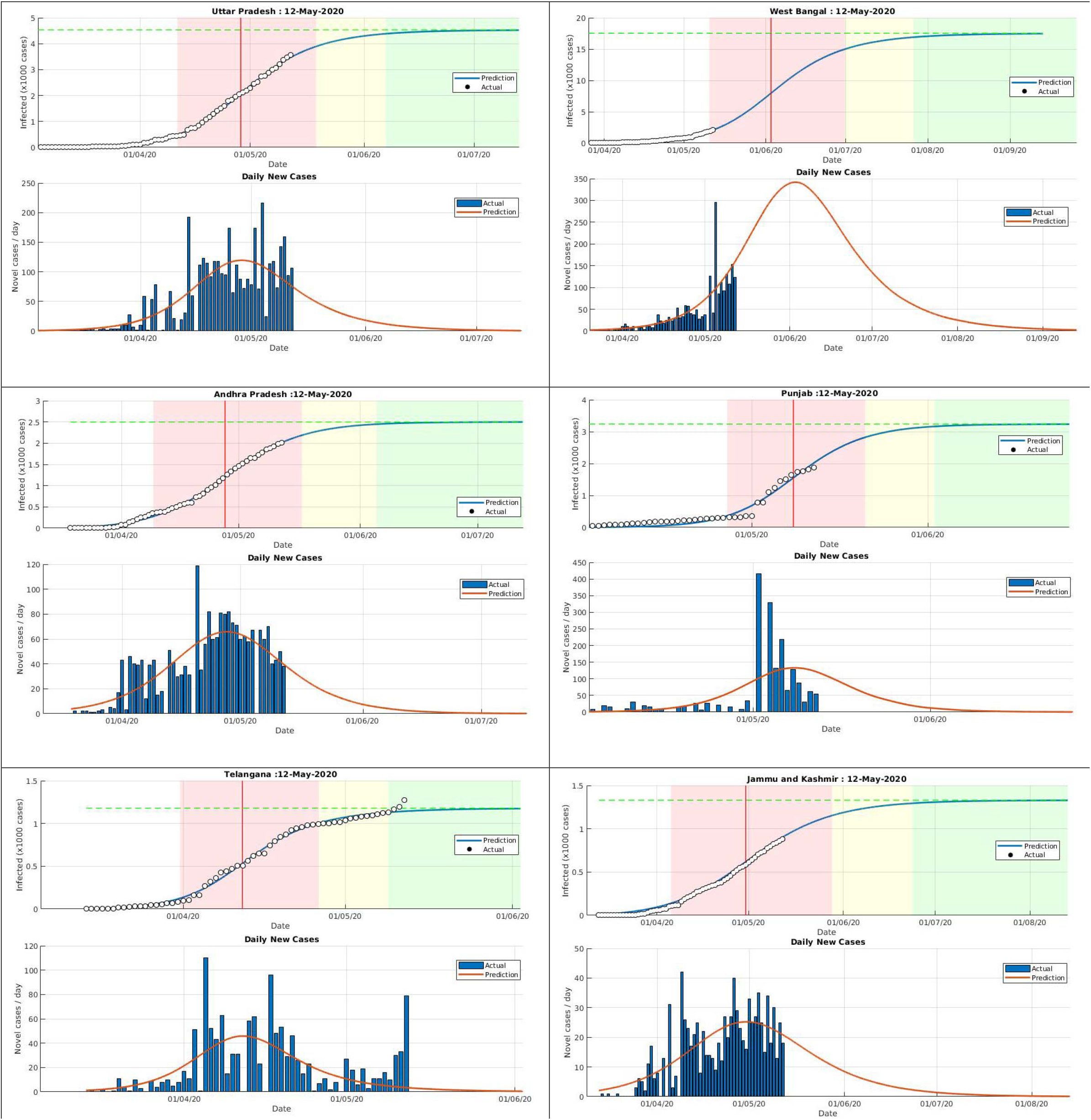
Predictions for various states (Uttar Pradesh, West Bangal, Andhra Pradesh, Punjab, Telangana, Jammu and Kashmir) using SIR model. The different epidemic phases are shown with white, red, yellow and green colors which represent initial exponential growth, fast growth (with positive and negative phase separated by red vertical line), asymptotic slow growth and curve flattening, respectively.

**Figure 4:**
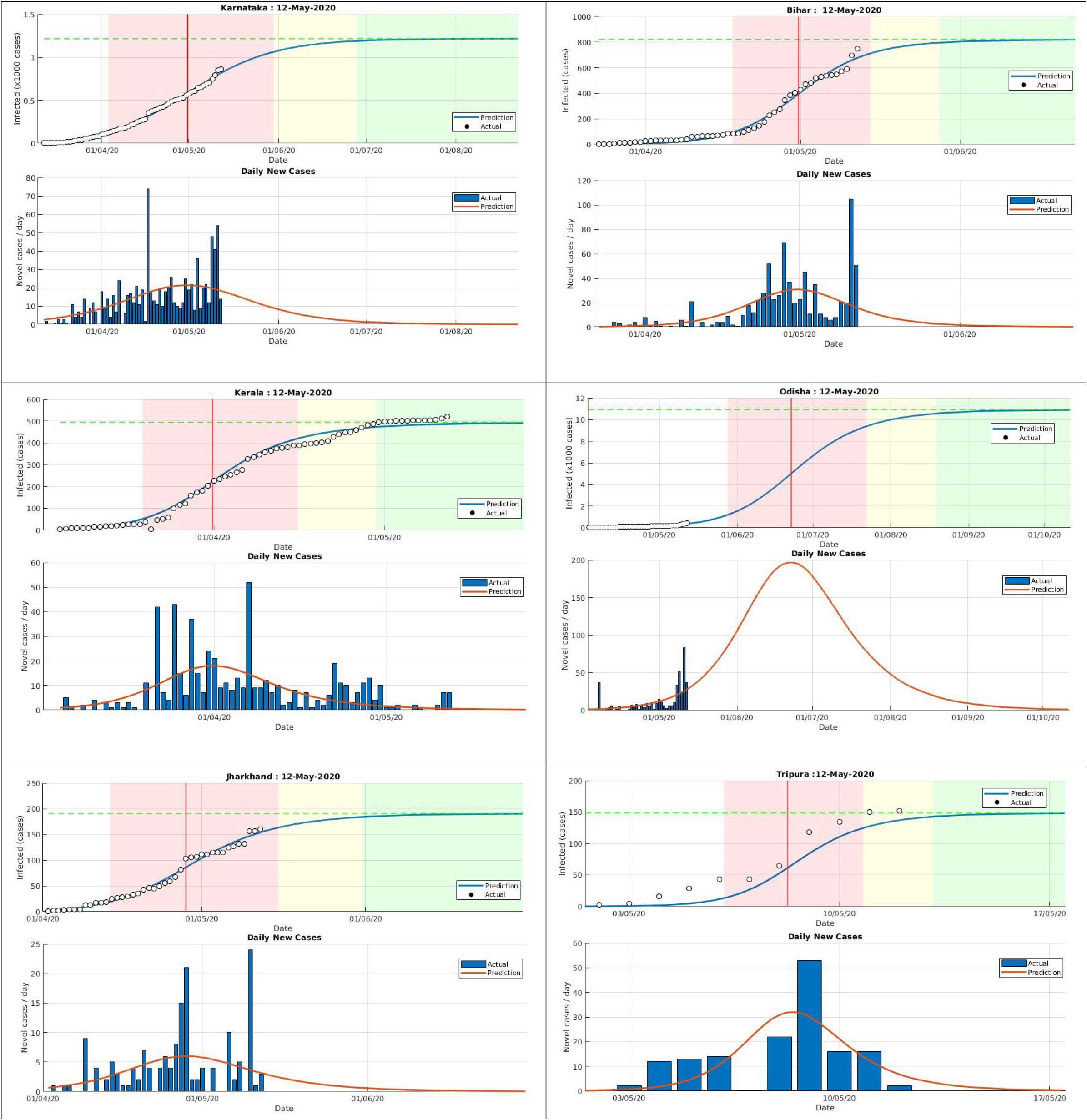
Predictions for various states (Karnatks, Bihar, Kerala, Odisha, Jharkhand and Tripura) using SIR model. The different epidemic phases are shown with white, red, yellow and green colors which represent initial exponential growth, fast growth (with positive and negative phase separated by red vertical line), asymptotic slow growth and curve flattening, respectively.

**Figure 5:**
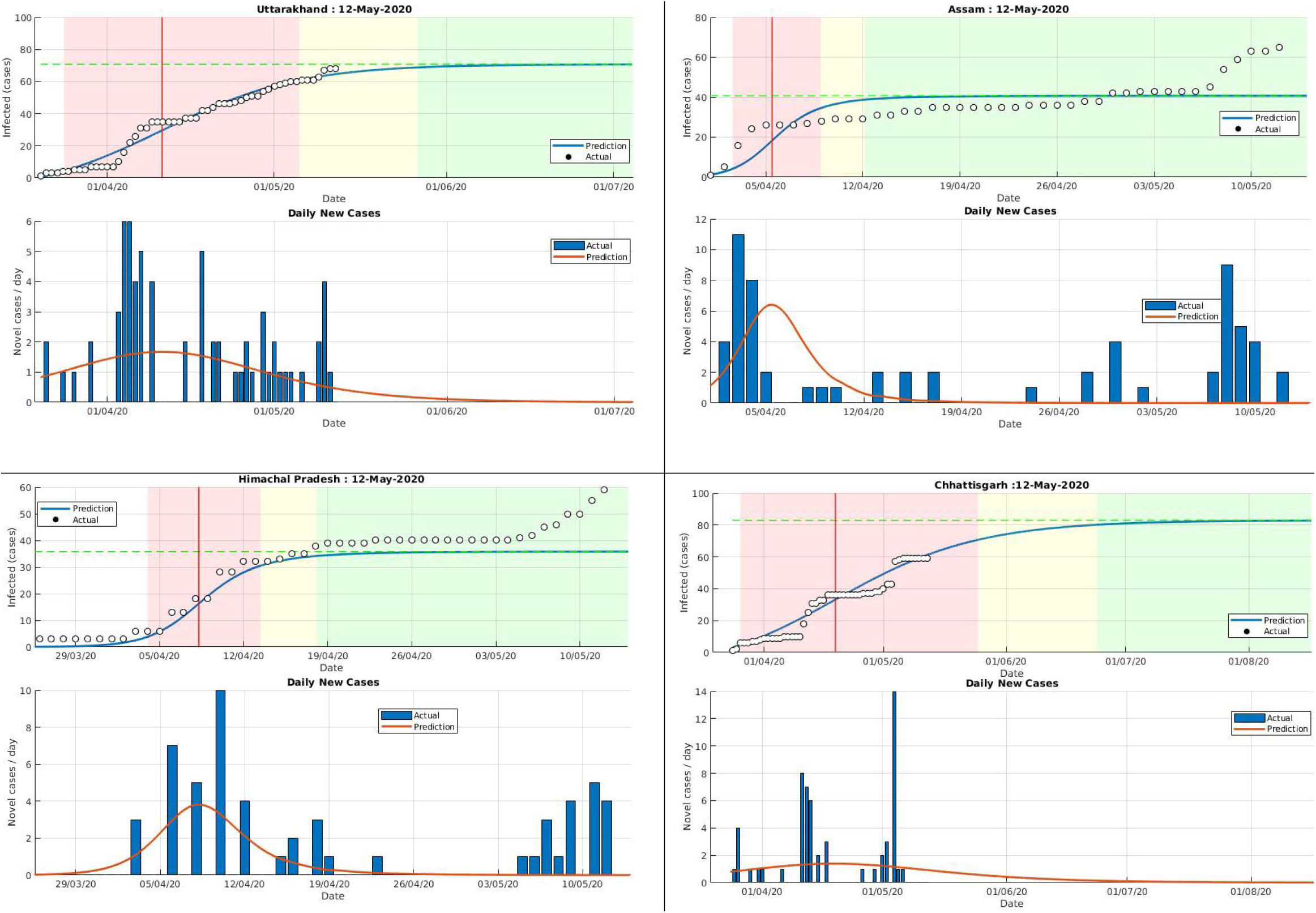
Predictions for various states (Uttarakhand Assam, Himachal Pradesh and Chhattisgarh) using SIR model. The different epidemic phases are shown with white, red, yellow and green colors which represent initial exponential growth, fast growth (with positive and negative phase separated by red vertical line), asymptotic slow growth and curve flattening, respectively.

**Table 2:** Statistics of SIR model for India and its various states.

| India and State | Number of observations | Degrees of freedom | RMSE | R<sup>2</sup> | Adjusted R-Squared | F-statistics vs. zero model | p-value |
| --- | --- | --- | --- | --- | --- | --- | --- |
| **India** | 71 | 67 | 924 | 0.998 | 0.998 | 10131.5 | 7.90173e-88 |
| **Maharashtra** | 63 | 59 | 243 | 0.999 | 0.999 | 14329.8 | 2.14263e-84 |
| **Gujrat** | 54 | 50 | 277 | 0.989 | 0.989 | 1610.48 | 1.03819e-49 |
| **Tamil Nadu** | 56 | 52 | 1084 | 0.703 | 0.679 | 51.9686 | 1.14244e-15 |
| **Delhi** | 71 | 67 | 147 | 0.995 | 0.995 | 4286.36 | 1.81163e-76 |
| **Rajasthan** | 71 | 67 | 79 | 0.996 | 0.996 | 5611.5 | 2.27489e-80 |
| **Madhya Pradesh** | 52 | 48 | 105 | 0.933 | 0.930 | 703.996 | 5.5109e-34 |
| **Uttar Pradesh** | 70 | 66 | 45 | 0.998 | 0.998 | 14179.8 | 1.22704e-92 |
| **West Bengal** | 55 | 51 | 40 | 0.995 | 0.995 | 3552.06 | 3.52659e-59 |
| **Andhra Pradesh** | 55 | 51 | 35 | 0.998 | 0.997 | 6742.15 | 2.99025e-66 |
| **Punjab** | 56 | 52 | 108 | 0.964 | 0.961 | 506.256 | 1.88912e-38 |
| **Telangana** | 60 | 56 | 34 | 0.995 | 0.994 | 3415.41 | 2.33263e-63 |
| **Jammu and Kashmir** | 61 | 57 | 29 | 0.99 | 0.99 | 1977.92 | 1.4714e-57 |
| **Karnataka** | 62 | 58 | 19 | 0.995 | 0.994 | 3599.05 | 7.83511e-66 |
| **Bihar** | 51 | 47 | 26 | 0.987 | 0.986 | 1228.3 | 1.24776e-44 |
| **Kerala** | 69 | 65 | 22 | 0.987 | 0.987 | 1635.79 | 3.90792e-61 |
| **Odisha** | 53 | 49 | 21 | 0.956 | 0.952 | 370.222 | 1.19745e-33 |
| **Jharkhand** | 41 | 37 | 7 | 0.985 | 0.984 | 827.057 | 6.07144e-34 |
| **Tripura** | 33 | 29 | 12 | 0.935 | 0.926 | 113.566 | 3.96801e-16 |
| **Uttarakhand** | 53 | 49 | 4 | 0.975 | 0.973 | 627.915 | 1.16588e-39 |
| **Assam** | 42 | 38 | 10 | 0.506 | 0.453 | 19.5462 | 7.88278e-08 |
| **Himachal Pradesh** | 52 | 48 | 7 | 0.868 | 0.856 | 100.349 | 1.09406e-20 |
| **Chhattisgarh** | 50 | 46 | 5 | 0.925 | 0.918 | 191.294 | 5.55392e-26 |

According to our prediction for India from data up to 12^th^ May 2020, the curve flattening of epidemic will start from first week of July and the epidemic may end by the third week of October (Figure 1). The calculated value of R_0_ is 1.15 and the epidemic size (number of infected peoples) is estimated to be 1,76,126. The coefficient of determination, R^2^ = 0.998, and p-value close to zero indicates the statistical significance of our predicted results from epidemiological data of India. Note that number of cumulative cases up to 1^st^ May 2020 in India was predicted to be ∼36,000 in recent study [21]. Also, the value of R_0_ is calculated as 1.50 [17] and 2.02 [27] in recent studies.

Among all the Indian states, Maharashtra is severely affected by COVID-19 epidemic. The SIR model indicates that the acceleration period of infection is from 25-Apr-2020 to 26-June-2020.The ending phase of epidemic may occur in the last week of June and it may be finished by the second week of September with epidemic size of 55,631(Figure 2). The calculated value of R_0_ is 1.11 with R^2^ and p-value close to 1 and 0, respectively. Note that number of cumulative cases up to 1^st^ May 2020 for Maharashtra was predicted to be ∼9,800 in recent study [21].

Another important state is Punjab, where according to media reports over 90,000 NRI returned to Punjab from abroad in the first quarter of the year 2020 [28]. The model indicates that the fast growth of infection is from 27^th^ April 2020 to 2^nd^ June 2020, thereafter, curve flattening will start and the epidemic is expected to finished by the first week of July (Figure 3). The estimated number of infected peoples is 3237. The calculated value of R_0_ is 1.28 which is little higher than country (1.15). The statistical parameter R^2^ is calculated to be 0.964 and pvalue is close to zero. Our study suggests that Punjab has relatively good control over the epidemic which may be due to the strict prevention measures taken during curfew imposed by Punjab government since 22^nd^ March 2020 in whole state.

The predicted data for other states is tabulated in Table 1 and plotted in Figures 2–5. The epidemic in Kerala is in its final phase and is expected to end by first week of June. Other states such as Telangana and Jharkhand are predicted to get rid of COVID-19 by the end of June. The states like Delhi, West Bengal, Odisha have to wait for the end of this epidemic up to mid-October, end of November and first week of December, respectively. The epidemic size of COVID-19 outbreak in Delhi, West Bengal, Gujrat, Tamil Nadu and Odisha can reach as large as 23,635, 17,522, 16,409, 12,555 and 10,921, respectively. The R_0_ for various states varies from 1.03 in Uttarakhand to 7.92 in Bihar. Note that in recent study, the daily infection rate (DIR) of COVID-19 for Maharashtra, Delhi Gujrat, Madhya Pradesh, Andhra Pradesh, Uttar Pradesh and West Bengal has shown exponential growth [20].

The calculated value of R^2^ for Assam (0.506) and Himachal Pradesh (0.868) is lower than the desirable value. This indicates a small correlation between the predicted and actual numbers of the epidemic, which is due to the small number and large fluctuation of epidemiological data. The epidemic in these states were expected to end in April, however, increased number of cases have been reported in last few days (Figure 5), which are attributed to the interstate movement of significant number of migrants in these states [29]. Considering the strict prevention measures, this rise in cases is momentarily and these states are expected to mitigate the epidemic in coming days.

It is worth discussing on the number of samples tested for COVID-19 cases around the country. As on 12 May 2020, a total number of 17,59,589 samples have been tested throughout the country out of which 74,329 samples were tested positive which comes out to be 4.22% of total collected samples [30]. The % of positive cases in Karnataka, Odisha, Jharkhand, Uttarakhand, Himachal Pradesh and Assam is < 1%. The total confirmed cases, total samples tested and % of positive cases are listed in Table 3. In Maharashtra, percentage of positive cases is highest with 10.98% followed by Gujrat (7.45%), Delhi (7.19%) and Madhya Pradesh (4.92%). The greater number of testing in these states may be required in coming days and the predicted results for these states may change in near future.

**Table 3:** Number of confirmed cases, number of samples tested and % of positive cases in India and various states as on 12^th^ May 2020 [30].

| India and State | Confirmed Cases | Samples Tested | % of Positive Cases from Tested Samples |
| --- | --- | --- | --- |
| India | 74329 | 1759579 | 4.224249 |
| Maharashtra | 24427 | 222284 | 10.9891 |
| Gujrat | 8904 | 119536 | 7.448802 |
| Tamil Nadu | 8718 | 266687 | 3.269001 |
| Delhi | 7639 | 106109 | 7.199201 |
| Rajasthan | 4126 | 185610 | 2.222941 |
| Madhya Pradesh | 3986 | 80885 | 4.927984 |
| Uttar Pradesh | 3664 | 140166 | 2.614043 |
| West Bengal | 2173 | 52622 | 4.129452 |
| Andhra Pradesh | 2089 | 191874 | 1.088735 |
| Punjab | 1914 | 43999 | 4.350099 |
| Telangana | 1326 | Not available | Not available |
| Jammu and Kashmir | 934 | 53726 | 1.738451 |
| Karnataka | 925 | 116533 | 0.793767 |
| Bihar | 879 | 37430 | 2.348384 |
| Haryana | 780 | 62377 | 1.250461 |
| Kerala | 525 | 38547 | 1.361974 |
| Odisha | 437 | 66057 | 0.66155 |
| Jharkhand | 172 | 22815 | 0.75389 |
| Tripura | 153 | 10344 | 1.479118 |
| Uttarakhand | 69 | 10471 | 0.658963 |
| Assam | 65 | 21791 | 0.298288 |
| Himachal Pradesh | 66 | 12224 | 0.539921 |
| Chhattisgarh | 59 | 27339 | 0.215809 |

## Conclusions

In summary, classical SIR epidemiological model has been used to predict the COVID-19 outbreak in India and its different states. The value of R_0_ is calculated to be 1.15 for Indian population. Among Indian states, Maharashtra is severely affected where the ending phase of epidemic may occur in the last week of June and it may be ended by the second week of September. The model indicates that the curve flattening in Punjab may start in the first week of June up to first week of July. The states like Delhi, West Bengal, Odisha have to wait for the end of this epidemic up to mid-October, end of November and first week of December, respectively. These predictions are based on the assumptions that the current preventive efforts will be continue. In Maharashtra, percentage of positive cases with respect to total sample collected is highest with 10.98% followed by Gujrat (7.45%), Delhi (7.19%) and Madhya Pradesh (4.92%), which are higher than the percentage of country (4.22%).

## Data Availability

No specific data reported.

## Acknowledgement

We acknowledge Prof. Milan Batista, University of Ljubljana, Slovenia for helpful discussion regarding technicalities of the method used in this study.

